# An ethnographic study of mobilization, basic mobility, physical activity, and exercise during acute hospitalization in patients following hip fracture surgery

**DOI:** 10.64898/2025.12.18.25342580

**Authors:** Maria Swennergren Hansen, Meta Ellen Staal Høite, Thomas Bandholm, Sofie Tscherning Lindholm, Kira Marie Skibdal, Mette Merete Pedersen, Jeanette Wassar Kirk

**Author notes:** CORRESPONDANCE: Maria Swennergren Hansen.

## Abstract

**Introduction:** Existing evidence highlight the need for a standardized approach in treating patients hospitalized following a hip fracture surgery. Clinical guidelines highlight rapid recovery of basic mobility post-surgery and the importance of frequent mobilization, physical activity, and exercise in rehabilitation. However, findings from the first HIP-ME-UP study show increasing physical inactivity among these patients, indicating issues with guideline implementation in clinical practice. Therefore, this study investigated clinical practice in relation to mobilization, basic mobility, physical activity and exercise during acute hospitalization in patients following hip fracture surgery.

**Methods:** An ethnographic field study was conducted at the orthopedic ward of a publicly funded hospital in the Capital Region of Denmark. Researchers carried out 22 observations from November 2023 to February 2024. A total of 230 pages of field notes were produced. An initial inductive qualitative content analysis was applied to find preliminary themes and subthemes. To nuance and validate these findings, 23 interviews with healthcare professionals and management were conducted and analyzed deductively. This deductive analysis was guided by the preliminary themes and subthemes that were revised in the final analysis if data provided nuances and new insights to the observations.

**Result:** Three themes emerged from the final analysis: (1) Different cultural models; (2) Organizational factors shaping mobilization practice; and (3) Mediating artefacts. Professional groups differ in their understanding of patient mobilization, leading to unclear responsibilities and inconsistent practices. A lack of interprofessional communication, resource constraints and challenging procedures for pain management aggravate these issues and hinder effective patient mobility. Moreover, the physical spaces and environments at the hospital contribute to patients staying in bed.

**Discussion:** Achieving recovery basic mobility and promoting mobilization, physical activity and exercise during hospitalization for patients following a hip fracture is complex and hindered by unclear definitions and responsibilities. The findings suggest a need for clear communication and consistent practices across professional and cultural boundaries to improve patient outcomes.

**Conclusion:** Our study contributes to an understanding of why mobilization remains inconsistently practiced despite guidelines that stress frequent mobilization as being important. These inconsistencies are deeply rooted in different cultural models regarding professional responsibility and practices, organizational priorities, and the hospital environment.

**HIGHLIGHTS:**

1. Cultural models shape how mobilization is defined, prioritized, and delegated in clinical orthopedic practice.
2. Professional roles and responsibilities around mobilization are negotiated through implicit routines and interactions.
3. Lack of a shared terminology regarding mobility limits interprofessional collaboration and contributes to fragmented mobilization practices.
4. Mediating artefacts like the Sara Stedy reflect and reinforce shifting norms about mobility in hospital care.
5. Organizational structures and staffing patterns influence how and when mobilization is implemented in daily routines.

## INTRODUCTION

The typical patient sustaining a hip fracture is approximately 80 years old (1), often frail (2) and living with multiple comorbidities (1). For these patients, a fall resulting in hip fracture is not only a major trauma but also a life-altering event (3,4). The subsequent surgery represents a second trauma, and the immediate postoperative period becomes a critical window that may determine long-term clinical outcomes (5). During this phase, prolonged immobility is associated with serious complications such as pulmonary disorders, hypotension, urinary retention, constipation, pressure ulcers, and increased risk of infections (6–9).

Previous studies have demonstrated that providing physiotherapy two (10) to three times (11,12) per day may improve clinical outcomes, including shorter length of stay (11,12) and a higher proportion of patients regaining basic mobility at discharge (10), compared to patients receiving physiotherapy once daily. In addition, associations have been seen between regaining pre-fracture independence in basic mobility activities before discharge and fewer readmissions, and lower mortality rates (13). However, translating guideline recommendations into clinical practice remains a challenge. Observational studies have shown that patients remain alarmingly inactive during hospitalization after hip fracture surgery (14), and that in-hospital activity levels have declined over the last decade (15,16). One contributing factor may be conceptual ambiguity and inconsistent terminology regarding mobility in clinical practice. Previous qualitative work in medical wards has shown that terms related to mobility are often used interchangeably or differently across professional groups, creating confusion and limiting coordination (17). This gap between evidence and practice has been described as a “knowing–doing gap” (18,19).

To address this gap, the HIP-ME-UP research program (***U****nderstanding and explaining clinical **p**ractice regarding basic **m**obility, physical activity and **e**xercise in patients following a **hip** fracture*) was established in 2022. The overarching purpose is to understand and explain clinical practices related to mobilization, basic mobility, physical activity, and exercise among patients hospitalized post-hip fracture surgery. While previous ethnographic studies have investigated mobility among older adults in medical wards (17,20), little is known about how mobility-related care unfolds in orthopedic settings following hip fracture surgery. This lack of insight is concerning, given the established links between early mobilization and reduced mortality (9,21) and better patient-reported outcomes (21). Therefore, the aim was to investigate clinical practice in relation to mobilization, basic mobility, physical activity and exercise during acute hospitalization in patients following hip fracture surgery. Clinical practice was defined by the research group as language, concepts, physical spaces and settings, actions and interactions between healthcare professionals and/or patients. All these factors together constitute local culture.

## METHODS

This qualitative study is situated within a critical realist framework (22,23), and drew on hermeneutic principles (24,25) to explore how organizational and institutional structures are interpreted, negotiated, and carried out by healthcare professionals in their everyday practice. In other words, critical realism guided our understanding of what structures exist and how they exert influence, while hermeneutics enabled us to investigate how these structures are experienced and given meaning. By integrating hermeneutic interpretation within a critical realist ontology, we conducted a layered analysis that addressed both the structural conditions shaping action and the interpretive processes through which action is understood. This combined lens is valuable in complex institutional settings such as healthcare, where structural conditions and health care professionals are in constant interaction.

Because terms related to mobility are often used interchangeably or differently across professional groups (17), and because they are central to the current study, we define them initially. We use *mobility* as an umbrella term that includes several related but distinct concepts: *mobilization* (assisted movement from one place to another, e.g. from bed to chair) (26), *physical activity* (any movement producing energy expenditure (27), and *exercise* (structured, therapeutic physical activity) (28). We also refer to *basic mobility*, as assessed by the Cumulated Ambulation Score (CAS) as it unfolds (29), and *early mobilization*, defined as initiating weightbearing within 24 hours post-surgery (30).

### Study design

#### Ethnography

To explore how clinical practices related to mobilization, basic mobility, physical activity, and exercise are carried out following hip fracture surgery, we conducted an ethnographic field study. This design was chosen to give us the opportunity to examine everyday practice as it unfolds within the social, material and institutional context of an acute orthopedic ward (31). Our ambition was not only to describe the culture related to mobilization, basic mobility, physical activity, and exercise, but to understand how it is embedded in daily practice: how it is understood, prioritized or delegated across different professions (32).

#### Semi-structured interviews

To enhance the depth and credibility of the analysis, and in line with principles of methodological triangulation (33), semi-structures interviews were conducted with professionals and departmental management. The interview guide was informed by a preliminary analysis of the field notes, enabling us to elaborate on emerging patterns, probe for alternative perspectives, and critically assess initial interpretations.

### Study setting

The study was conducted at a publicly funded hospital in the Capital Region of Denmark, part of the tax-financed Danish healthcare system. Data were gathered across two orthopedic wards. Ward A (32 beds) and Ward B (26 beds) that both admit patients with a range of traumatic orthopedic injuries. The clinical staff in both wards include nurses, nurse assistants, orthopedic surgeons, and pharmacists. Physiotherapy and occupational therapy services are provided by a centrally organized rehabilitation department.

Physiotherapists are typically assigned consistently to the orthopedic wards, allowing for continuity in daily clinical practice, whereas occupational therapy is delivered by a rotating team of six therapists serving the entire hospital. Both professions aim for continuity, with the same therapist following the patient throughout hospitalization, but this varies due to shift schedules, task distribution, and weekday coverage. This organizational structure results in variability in both staff presence and treatment options available to patients on a day-to-day basis.

Situated within a critical realist paradigm, this description of the study setting reflects an understanding that clinical environments are shaped by both observable practices and the underlying organizational mechanisms that structure professional roles, resource allocation, and care delivery. Acknowledging these contextual dynamics is essential for interpreting the actions and experiences of healthcare professionals within the specific institutional conditions in which they operate.

### Data collection

#### Ethnography

Participant observations were carried out between November 2023 to February 2024. Observations focused broadly on patient–staff interactions, with particular attention, when possible, to individuals recovering from hip fracture surgery. Participant observations were primarily conducted by the last author (JWK), a registered nurse, with extensive clinical and ethnographic experience, and a research assistant (MS) trained in public health science and possessing more limited clinical and ethnographic experience.

Their different levels of experience provided complementary observational perspectives and preunderstandings. This enabled two complementary observation strategies: MS followed one health professional at a time, while JWK adopted a more dynamic approach, shifting focus based on emerging insights and alignment with the research aims. To incorporate a physiotherapeutic perspective, a physiotherapist (MMP) conducted three additional observations focusing specifically on physiotherapy practices in Wards A and B. In total, 22 field observations were carried out.

The initial HIP-ME-UP study identified 10:00h –12:00h as the most physically active period of the day for patients (15). Therefore, most observations were scheduled within this timeframe. To gain a broader understanding, two additional observations were conducted during the evening shift.

The participant observations were conducted in a primarily non-participant position (34), a deliberate distance to focus on documenting clinical practices and social interactions without actively engaging in them. This stance allowed for close attention to the unfolding of everyday activities while minimizing interference in the dynamics of the observed setting. The researchers occasionally engaged in brief clarifying dialogues with staff members; however, more in-depth inquiries were reserved for follow-up interviews.

The observation guide was inspired by Spradley’s framework, which conceptualizes social situations as composed of three elements: person, activity, and place (31). Field notes contained both descriptive and analytical content, capturing contextual details alongside reflections of underlying structures shaping clinical practice. All notes were elaborated upon immediately following each observation, resulting in approximately 230 pages of field notes.

To enhance the credibility (35) of the findings, two consensus meetings were held during the data collection period. All authors participated in these meetings with the aim of refining the observational focus, ensuring methodological consistency, and discussing emerging patterns. Prior to the second meeting, JWK, MS, and MSH conducted an initial review of all field notes and discussed the degree of data saturation. This process led to the identification of three underexplored areas for further observations: communication between surgeons and patients, interactions involving occupational therapists, and physiotherapy sessions conducted in the afternoon. As a result, data collection was extended by three additional days to explore these areas more thoroughly.

#### Semi-structured interviews

The interviews were conducted in September 2024. We conducted 23 interviews with healthcare professionals (N=18) and managers (N=5). The interviews were conducted by MSH, JWK, KMS, and MMP, all with clinical backgrounds and varying experience in qualitative interviewing. The interviews were conducted either in offices at the wards, in meeting rooms at the physiotherapy therapy department, or in the managers’ offices. The interviews lasted around 30-40 minutes each. All interviews were audio recorded and transcribed verbatim, resulting in 280 pages of transcript data.

### Participants

Participants in the field observations were healthcare professionals working in the orthopedic wards. In this study, the term *healthcare professionals* refer to all staff groups involved in patient mobilization, physical activity and exercise, including physiotherapists, occupational therapists, nurses, nursing assistants, porters, and orthopedic surgeons. The term *nursing staff* includes both registered nurses and nursing assistants. Participants in the interviews included both healthcare professionals and members of the management team (Table 1). A purposive sampling strategy (36) was employed to ensure variation in professional roles and perspectives relevant to mobilization, basic mobility, physical activity and exercise practices.

**Table 1.**
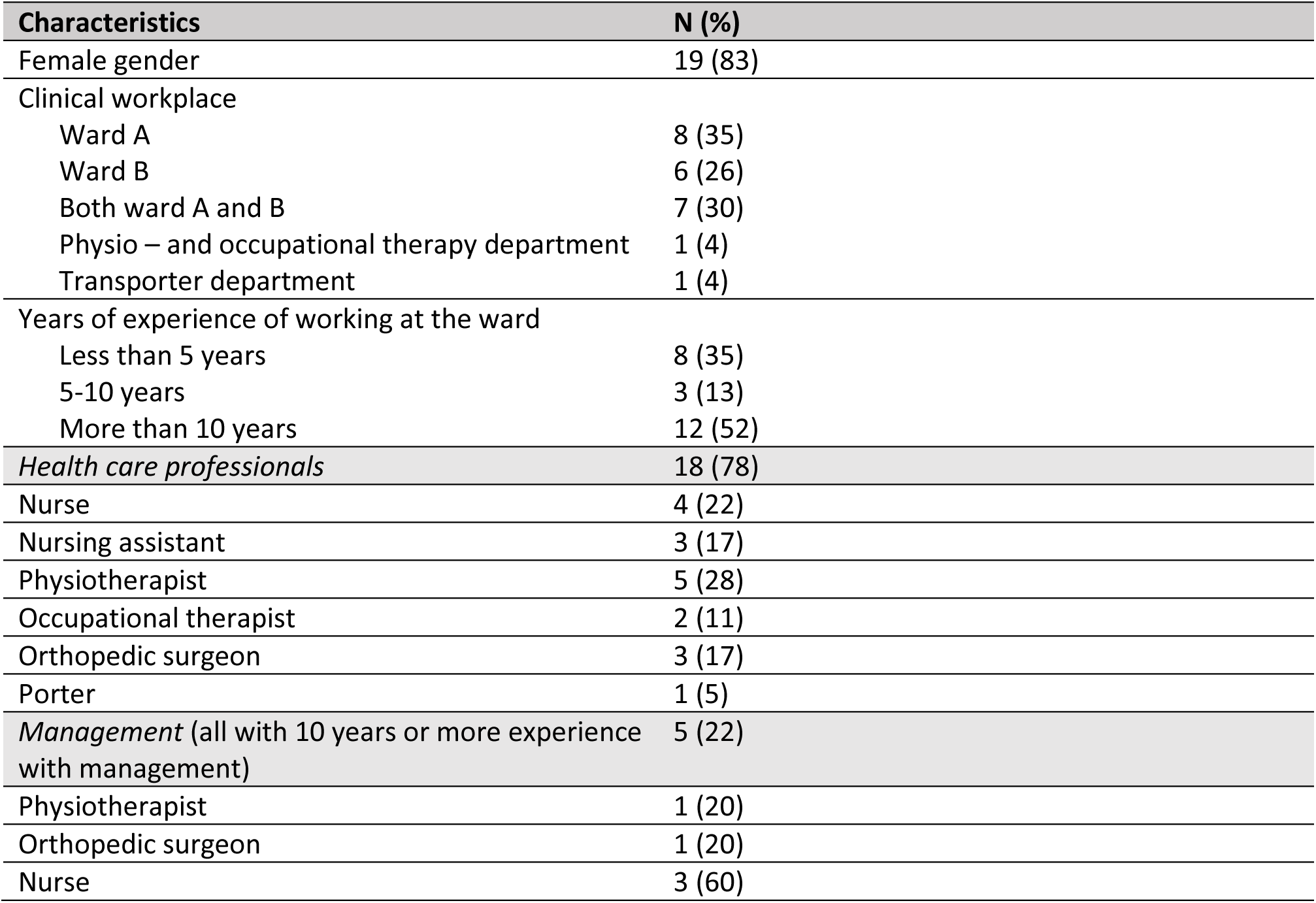
Participant demographics interviews (n= 23).

| Characteristics | N (%) |
| --- | --- |
| Female gender | 19 (83) |
| Clinical workplace |  |
| Ward A | 8 (35) |
| Ward B | 6 (26) |
| Both ward A and B | 7 (30) |
| Physio – and occupational therapy department | 1 (4) |
| Transporter department | 1 (4) |
| Years of experience of working at the ward |  |
| Less than 5 years | 8 (35) |
| 5-10 years | 3 (13) |
| More than 10 years | 12 (52) |
| *Health care professionals* | 18 (78) |
| Nurse | 4 (22) |
| Nursing assistant | 3 (17) |
| Physiotherapist | 5 (28) |
| Occupational therapist | 2 (11) |
| Orthopedic surgeon | 3 (17) |
| Porter | 1 (5) |
| *Management (all with 10 years or more experience with management)* | 5 (22) |
| Physiotherapist | 1 (20) |
| Orthopedic surgeon | 1 (20) |
| Nurse | 3 (60) |

### Ethics

The study was approved by The Research Ethics Committee of the Capital Region of Denmark (journal number: F-23067628, 8/11/2023) and conducted in accordance with the Declaration of Helsinki (37). Additional institutional approval was granted by senior management in both the Orthopedic and Physio – and occupational therapy Departments, as well as by the head nurses of the two participating wards.

However, verbal consent was granted before following health care professionals on a one-to-one basis. Patients were not the focus of observation and did not interact directly with the researchers; all fieldnotes were anonymized, and no identifiable patient data were recorded.

A *situated ethics* (35,38) approach was adopted, acknowledging that ethical practice in qualitative research must be responsive to context and relational dynamics. This approach was particularly relevant within the complexity of clinical environments, where ethical decisions are continuously negotiated in situ.

Throughout the fieldwork, the observers engaged in ongoing reflexivity, adapting their presence and degree of engagement in response to evolving clinical circumstances to minimize disruption and ensure respect for both staff and patients.

In line with a critical realist perspective (22,23), we approached ethics not only as a matter of formal procedures but as something embedded in everyday clinical work. This meant recognizing that healthcare practice is shaped by both individual choices and broader institutional and social conditions. In practical terms, the researcher acted to minimize intrusion during sensitive situations—for example, by stepping back or leaving the room when private patient matters were discussed during ward rounds. To protect confidentiality, field notes were anonymized and contained no personal identifiers. Before each interview, participants received both verbal and written information about the study, and written informed consent was obtained.

The study was pre-registered at OSF (https://doi.org/10.17605/OSF.IO/3FN59). As the study progressed, the analytical strategy was refined in response to insights emerging from the empirical data (field notes) consistent with the overall approach for data analysis specified in the pre-registration. In reporting this study, we adhered to the Standards for Reporting Qualitative Research (SRQR) (33) and the REPORT trial guide (39).

### Data analysis

The analysis followed a two-step approach, beginning with an inductive analysis of the field notes and followed by a deductive analysis of the transcribed interview data (40).

During the data collection period, MSH engaged in ongoing reading of the field notes to trace the development of emerging insight and to assess the progression toward data saturation. Subsequently, each field note was re-read in detail and divided into meaning units that were condensed at the manifest level. Where relevant, latent codes were also noted either during this process or through iterative re-reading of the material. Analytical memos were written throughout to capture emerging analytical questions and guide the formulation of topics for further exploration in the interviews.

To enhance credibility (41) of the analysis, selected samples of the coding and condensation process were cross-checked by JWK, who also contributed to the development of the thematic abstraction (42). Based on the coded and condensed meaning units, MSH constructed an initial thematic structure, grouping related condensates into subthemes and overarching themes (41). This structure was discussed and revised during a meeting (attended by MSH, STL, KMS, JWK), reducing the number of main themes from seven to five. The final theme names and structures were refined by MSH, with full consensus achieved during an author meeting, to ensure analytical trustworthiness.

Although the primary focus was on patients with hip fractures, all observed clinical interactions were included in the analysis, based on the critical realist understanding that all patient-staff interactions contribute to the cultural and organizational context of the ward.

The interviews were transcribed using AI-assisted software and manually verified for accuracy by MSH or STL. The deductive analysis began with full reading of all interview transcripts (278 A4 pages) by MSH to establish familiarity with the data material. A systematic text condensation process was then applied to each transcript, and meaning units were identified, condensed, and coded in relation to the preliminary themes. Quotations that were representative of the subthemes were noted for inclusion in the results. Special attention was given to data that confirmed, nuanced, or challenged insight from the field observations. Revisions to themes and subthemes were made in dialogue with STL and JWK. The final analysis was discussed and consolidated in a full author meeting (excluding MS).

## RESULTS

The analysis showed that getting patients out of bed and moving following a hip fracture surgery is a complex task where several factors on many different levels affect clinical practice. This is described in terms of three themes (**Fig 1**.): (1) Different cultural models; (2) Organizational factors shaping mobilization practice; and (3) Mediating artefacts. The themes include 9 subthemes that elaborate on the complexities of in-hospital patient mobility following hip fracture surgery. These themes and subthemes will be presented in the following section.

**Fig. 1.**
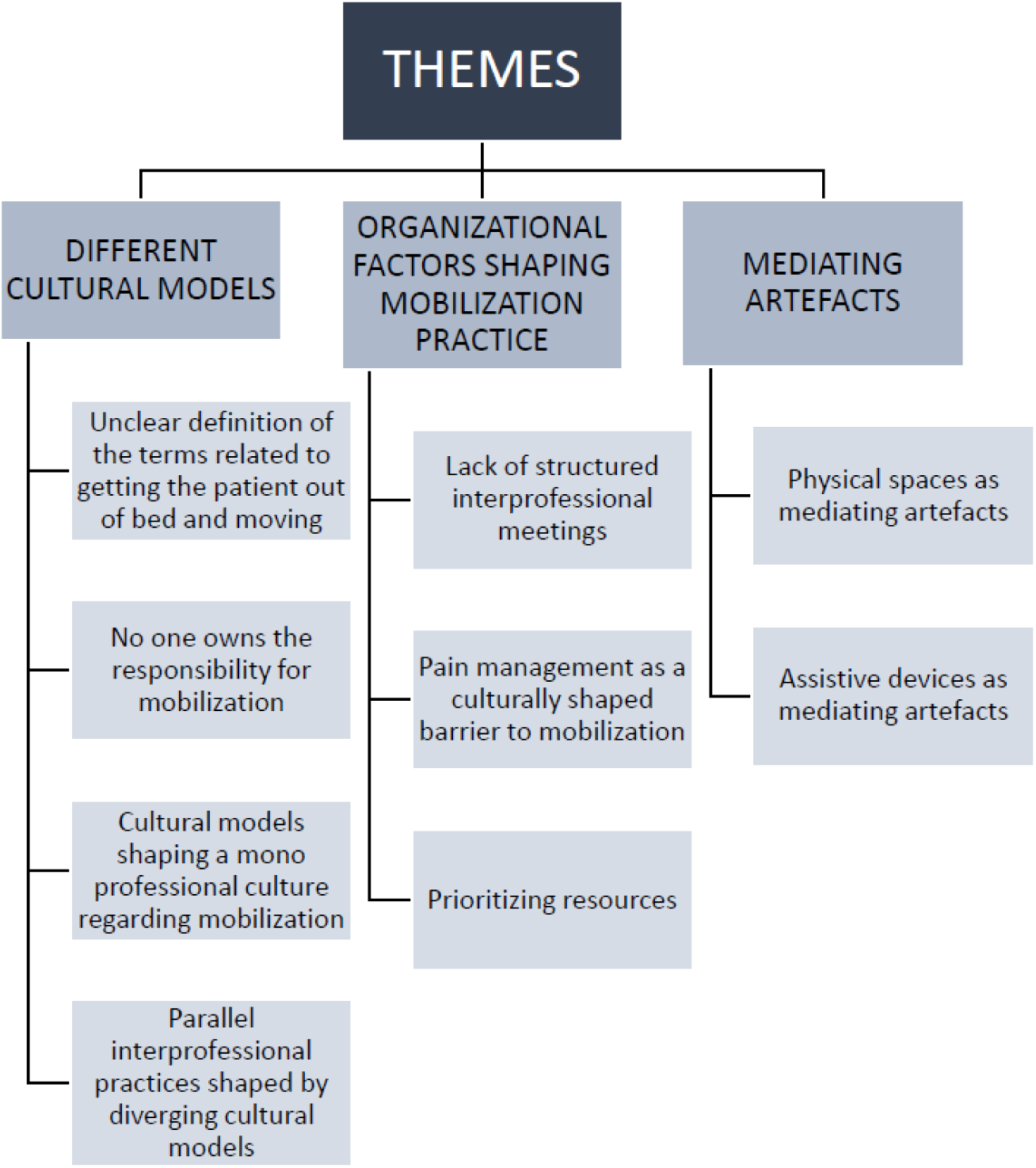
Themes and subthemes.

### Different cultural models

This theme captures how different professional groups e.g. nurses and therapists draw on distinct cultural models in their understanding and handling of patient mobilization, physical activity and exercise. *Cultural models*, as conceptualized by Hasse (43), refer to the shared and taken-for-granted knowledge that guides how we think, speak, and act within specific social contexts. These models operate as cultural logics — simplified and organized cultural connections that shape our expectations about how the world works and how certain tasks should be performed. Cultural models do not simply exist; they are produced and reinforced through daily interactions, routines, and the use of specific artefacts. They emerge as novices observe and imitate experienced colleagues, as team members coordinate care, and as particular ways of talking and doing become habitual over time. In this process, institutional structures, professional hierarchies, and embedded power relations act as formative forces, privileging certain interpretations and practices while marginalizing others. A physiotherapist, for example, may learn through repeated participation in monodisciplinary meetings where the physiotherapeutic perspective and way of handling mobilization is highlighted. Over time, these patterned understandings crystallize into shared assumptions that require no explicit explanation among insiders. A concrete example of how such implicit assumptions is communicated can be seen in the routine practice among physiotherapists of always closing the door to the local therapy room, which also serves as a space for medical record documentation. When a newcomer leaves the door open, a more experienced staff member casually remarks, “*Remember to close the door*.

*We usually do that*” (fieldnotes). This seemingly minor correction reflects an unspoken norm that has been internalized through repeated enactment, illustrating how cultural models are sustained through small, everyday interactions. At the same time, the closed door may signal to patients that the room functions primarily as an office rather than a space intended for exercise or physical activity.

The analysis shows that the dominance of certain cultural models in the clinical context often depends on organizational positioning or professional status. Actions that deviate from these cultural models are quickly noticed and sometimes resisted, as they challenge what is perceived as proper or expected practice. The following subthemes illustrate how these divergent cultural models influence collaboration, task allocation, and perceptions of patient mobility in everyday clinical work.

### Unclear definition of the terms related to getting the patient out of bed and moving

During the observations, it became evident that there is no shared language for the task of “getting the patient out of bed and moving.” Nursing staff most frequently use the term mobilization, which they understand as assisting patients in moving from the bed to a chair or to the toilet. Mobilization is considered part of routine care, typically linked to tasks such as personal hygiene or nutrition—core elements of nursing work. Therapists, on the other hand, also use the term exercise. For physiotherapists, mobilization tends to refer to everyday activities, whereas exercise is understood as structured physical training prescribed with a specific therapeutic purpose, following principles of frequency, intensity, and duration. The term physical activity does not appear in the vocabulary of nursing staff during clinical interactions. While physiotherapists refer to physical activity when discussing overall treatment goals, it is rarely used in day-to-day communication.

The observations revealed that the meaning and use of the terms mobilization, physical activity, and exercise differ by profession and are shaped by distinct cultural models. These differing understandings influence who takes responsibility for the task, how it is carried out, and with what intended purpose.

### No one owns the responsibility for mobilization

As one physiotherapist put it: “*Every man and no man’s responsibility*” (Physiotherapist 3). During the observations it became clear that the responsibility for mobilization following hip fracture surgery is diffuse, with differing views across professional groups and thereby cultural model. Observations indicate that nursing staff primarily view mobilization as a part of nurses and assistants’ duties. However, many expressed feeling overwhelmed by competing tasks that contribute to a tendency to share or even relinquish this responsibility. As stated by a nurse:

> *"We mobilize to the extent that we can. All our patients need to be mobilized. But there aren’t always enough of us to assist everyone multiple times a day. Therefore, our efforts are limited."* (Nurse 5).

During observations, nurses expressed feeling caught between their many caregiving tasks:

> *“In reality, it doesn’t take that long. But it does when you have a lot of tasks to manage, and this isn’t your only area of focus”* (Nurse 12).

On the other hand, physiotherapists view mobilization as a shared responsibility up for debate, noting that nurses often expect physiotherapists to handle it. One physiotherapist commented:

> "*Mobilization we also do, but if it’s just for the purpose of getting them from A to B, then I would say they have to ask a porter to do it or do it themselves. If it’s mobilization with purpose, then yes, you’re involved"* (Physiotherapist 20).

Surgeons at the ward were seldom observed engaging in mobilization. They define weightbearing limits, set guidelines, and encourage patient adherence, but do not participate in the practical aspects.

Due to the limited number of occupational therapists employed at the hospital, their presence during observations was minimal. They view themselves as part of the mobilization task, particularly for patients requiring greater assistance, and sometimes this includes helping patients move from point A to B as part of their assessment. They expressed the belief that mobilization is a shared responsibility across professions. However, their main focus is on activities of daily living (ADL).

> “*We are a bit more involved in training. Specifically, we focus on ADL training. We also help mobilize the heavier patients who need assistance to do so*” (Occupational therapist 18)

### Cultural models shaping a mono professional culture regarding mobilization

From a cultural model perspective, the prioritization of mobilization tasks reflects deeply embedded understandings of professional roles, responsibilities, and identities that are stabilized through everyday practice. These cultural models are not always made explicit, but they guide how different professionals act, what they consider important, and how they interpret their roles in relation to others.

This mono professional culture is further evident in how mobilization is organized and practiced. Physiotherapists are culturally positioned as the primary healthcare professionals responsible for ensuring compliance with national clinical standards, such as the requirement that patient bear weight on their legs within 24 hours after hip fracture surgery (1). Although such documentation could, in theory, be completed by any professional, cultural models position this responsibility within the physiotherapy domain. This is reflected in practice, where physiotherapists not only carry out the assessments but also feel an implicit responsibility to be the first to mobilize patients:

> *“Nine out of ten patients (with a hip fracture), we’re the ones who get them up for the first time. It’s somewhat implicit up there, even if it’s not written down. But for other patients—for example, amputation patients or those with other fractures—it doesn’t necessarily have to be us who mobilize them first” (*Physiotherapist 22).

These implicit understandings, also create tensions in interprofessional collaboration. The nursing staff perceive that the early mobilization agenda is strongly driven by physiotherapy priorities which undermine shared responsibility. At the same time, physiotherapists may operate with a cultural model that frames their role as assessors rather than trainers, leading to further mismatches in expectations. As one nurse put it:

> *"Physiotherapists became highly focused on identifying which new patients needed to be assessed. And they do this assessment task, often with a porter or someone. And once they’ve made their assessment, their job is like finished"* (Nurse 1).

Finally, interviews revealed that mobilization, despite its clinical importance, is vulnerable to being deprioritized in practice. Reduced staffing and shifting generational attitudes among nurses contribute to changing cultural models of what constitutes essential care. As one nurse noted: “*When there isn’t a necessary task with a patient, mobilization is what gets sidelined*” (Nurse 1).

A more experienced nursing assistant reflected on generational differences:

> “*Those of us who’ve been here many years are more eager to ensure that patients are mobilized. It seems like it is not as important to the new generation*” (Nursing assistant 10).

### Parallel interprofessional practices shaped by diverging cultural models

Our empirical findings shows that collaboration around patient mobilization after hip fracture unfolds along a continuum with three levels; 1) mono professional practice, where work is carried out entirely within one profession; 2) interprofessional practice where professions coordinate their work while maintaining clear role boundaries; and 3) cross professional practice, the most integrated form, where tasks and responsibilities are shared or transferred across professions.

Our findings suggest that mobility practice in the studied wards rarely reaches the cross professional level. Instead, it often takes the form of parallel interprofessional practice, a variant of interprofessional work in which different professions operate side by side on the same patients but with limited integration of tasks or shared decision-making. This pattern is shaped by diverging cultural models of roles, responsibilities, and priorities, which influence how tasks are discussed, organized, and performed.

Although interprofessional dialog about patients and their treatment does occur, it rarely brings mutual insight into each other’s work. A senior surgeon described good collaboration with therapists based on long-standing familiarity but doubted that newer colleagues had the same experience, underscoring how shared cultural models can depend on long-term relationships:

> “*If you don’t know each other, it’s not as easy to initiate collaboration—especially when it comes to tasks at the interface between roles. Not knowing each other reinforces silo thinking*” (Surgeon 9).

A physiotherapist linked reduced cohesion to higher staff turnover:

> “*Today it’s not as organized, and people no longer know much about what others do. There’s also more turnover among the nursing staff, which makes it harder”* (Physiotherapist 19).

These statements reflect how the breakdown of shared cultural models weakens the basis for coordinated, cross professional work.

This divergence becomes particularly clear in relation to the mobilization task. Negotiations about who should perform it and how it should be done are ongoing, with some staff framing it as an “us vs. them” issue. A nurse remarked: “*If we need help…*” (Nurse 2) and “*If physio is there, then we can give a hand*” (Nurse 2). Similarly, a nursing assistant described the challenge of mobilizing a patient who requires two people while referring to nursing colleagues as “our own” rather than therapists, reinforcing the cultural boundary between groups :

> “*Because we can’t just pull in two of our own staff, as there simply isn’t enough personnel*” (Nursing assistant 10).

Within these cultural models, collaboration partners are often imagined within one’s own profession. Nurses tend to refer to other nurses or porters, not therapists when discussing teamwork. While good interprofessional collaboration was observed, it appeared dependent on individual relationships rather than on shared cultural models embedded in departmental routines. As stated by a physiotherapist:

> “*It’s very rare that we manage to carry out a joint intervention with the nursing staff. It’s like—one, where are they? Two, they withdraw. You can’t find them. It’s as if it’s not their responsibility. I keep searching and searching. That’s all I do. It’s really demotivating*.” (Physiotherapist 23).

In summary, these findings illustrate how distinct and sometimes competing cultural models across professional groups influence mobilization practices. These models are not fixed but are continually negotiated and reproduced through everyday interactions and institutional structures. As a result, interprofessional collaboration often unfolds in parallel rather than integrated forms, limiting the potential for shared responsibility and coordinated mobilization efforts.

### Organizational factors shaping mobilization practice

During observations, several organizational-level factors were found to influence how mobilization is carried out in the wards. These include the absence of a fixed interprofessional meeting structure, suboptimal coordination of pain management, and challenges related to resource prioritization.

### Lack of structured interprofessional meetings

The lack of formalized interprofessional meeting structures reinforces the parallel working practices described in the previous subtheme. During observations, the different professional groups rarely engaged in systematic or recurring collaborative forums. Instead, communication largely occured through a fragmented series of brief, task-oriented interactions

Typically, the nurse responsible for ward rounds gathers information from the night shift, reviews patient charts, and conducts a short morning meeting with the physio- and occupational therapists prior to the surgeon-led round. However, these meetings are highly compressed, with numerous patients needing to be addressed in a limited timeframe

Several nurses expressed that the morning meetings are too brief to cover both patients’ home situations and to engage in meaningful dialogue with physiotherapists to support planning of treatment. Nurses perceived that management encouraged efficiency over detail in these meetings. Physiotherapists, in turn, described the meetings as offering varying value, highlighting the challenge nurses faced in overseeing over 30 patients, many of whom they did not know well. A physiotherapist described the meetings as vague, lacking in specificity from both therapists and nurses:

> *“Sparing. I have to say that. It’s supposed to be a dialogue about the patient’s health status and their discharge plans in relation to the municipality and that sort of thing. But sometimes, it ends up being about small, insignificant matters. You really must be quite firm at times to push through an assessment related to discharge. Yes, both professions need to be better at being more specific about what we’re supposed to cover during these morning meetings. Sometimes it just turns into meaningless chatter” (*Physiotherapist 23).

This vagueness in interprofessional dialogue illustrates the presence of competing cultural models that shape how mobilization is understood across professions. These models, embedded in organizational structures, may inadvertently limit opportunities for coordinated decision-making and shared planning, as each profession draws on its own implicit assumptions and practices when engaging in dialogue.

There was also concern that information shared at the morning meetings were not always passed on to the nursing staff or the surgeons on rounds. For instance, situations were observed in which physiotherapists raised questions about weight-bearing status in newly operated patients—questions that the ward nurse could not answer. This uncertainty could potentially delay mobilization for that day.

Physio- and occupational therapists also reported a lack of knowledge exchange with both surgeons and pharmacists.

> “*One might think that the optimal solution would be to have a doctor present. There are probably some workflows that could move a bit faster” (Physiotherapist 17)*.

Although physio- and occupational therapists were, in principle, welcome to consult the surgeons during the day, but they often felt they were intruding.

“*If we’re in doubt about something or have questions, we go and grab them. It’s great that we can do that, but we’re interrupting them. They’re usually receptive to it, most of the time. But it is a disruption—interrupting them in the middle of something” (*Physiotherapist 19).

This illustrates how the absence of a structured interdisciplinary meeting negatively affects communication among staff, with direct consequences for patient mobilization.

### Pain management as a culturally shaped barrier to mobilization

Pain management is not merely a biomedical prerequisite for mobilization but is embedded in professional assumptions about roles, responsibilities, and patient agency. These implicit models influence how pain management is organized, who is expected to initiate it, and how it becomes integrated or overlooked, in the mobilization process.

Physiotherapists and nurses described postoperative pain management as suboptimal, with coordination gaps that delayed or prevented mobilization. Although most patients receive epidural analgesia during the first three postoperative days, staff noted instances where the non-operated leg became anesthetized, limiting safe mobilization until the epidural was adjusted or removed. Transitions from epidural to oral analgesics were described as error-prone, particularly when medications were not documented in the medical record and thus not administered.

Within the prevailing cultural model among surgeons and nurses in the ward, patients are expected to request supplementary analgesics themselves. This expectation reflects a cultural model in form of a tacit, shared understanding that positions patients as responsible agents in pain management, despite their condition. Such a model structures everyday practices, often invisibly, by defining what is seen as natural or appropriate. Several physiotherapists questioned the effectiveness of this approach, noting that many patients either refrain from asking or believe their requests will not be acted upon. In practice, this means that physiotherapists often delay sessions while searching for a nurse to administer extra medication:

> “*I don’t know where to begin. There’s so much communication, and you’re looking for the nursing staff—who aren’t there. And the pharmacists are only available for a certain period to help*” (Physiotherapist 23).

Nursing staff also acknowledged that attentiveness to pain can be inconsistent, reflecting competing priorities and embedded task hierarchies:

> *“But it absolutely also has to do with pain relief. And that’s on us. We can hear it at the morning meetings—it’s not always that we, as nursing staff, are attentive to when our patients are in pain, which prevents physios from mobilizing them”* (Nurse 2).

These accounts illustrate how shared, taken-for-granted assumptions about roles and responsibilities both enable and constrain mobilization practices. The model frames pain relief as a nursing-led task yet simultaneously shifts initiative to patients, revealing a contradiction between cultural expectations and clinical realities.

### Prioritizing resources

By observing physical artifacts on the ward, such as call bells ringing and patients seeking attention, it became clear that patients were actively trying to make contact. When the low level of mobilization was discussed during observations, most staff attributed it to a lack of resources. “Too few hands,” especially on evening shifts, as noted by nursing assistant 10. A nurse remarked:

> “*It’s very much about being busy. It’s not because we don’t want to. There’s simply no time. It sounds like a cliché, but that’s the narrative throughout the healthcare system. It’s not how it should be, but when you’re forced to prioritize in daily practice, this is often what gets deprioritized. Just like when you’re quick to call a porter because you’re in a rush—it’s just easier*” (Nurse 11).

Physiotherapists emphasized that many patients are physically heavy and require two people for mobilization. As no porter is permanently assigned to the ward and must be requested via a centrally managed app, physiotherapists are less likely to call on porters for assistance. Consequently, physiotherapists often mobilize patients in pairs, which naturally limits the number of patients they can treat in a day.

In terms of resource allocation across the day, physiotherapists are present on the wards during daytime hours (08:00h–15:30h). As shown in our previous study (15), most physical activity was observed around 11:00h – 12:00h —coinciding with peak staffing and physiotherapists’ “prime time.” Two nurses explained that few patients are mobilized during evening shifts due to staff shortages; activity is typically limited to toilet visits:

> “*After 3 p.m., no one really comes in anymore—only if patients use the call bell. I think they feel left to themselves*” (Nurse 13). “*Between 1 and 2 p.m., we don’t go into patient rooms unless they call for something, like needing to use the toilet. And on evening shifts, we’re so short-staffed that mobilization simply isn’t prioritized. That’s why we make sure they’re at least mobilized in the morning*” (Nurse 13).

From the patients’ perspective, mornings are marked by intense activity. A physiotherapist described the ward in the morning as “*like a central station—blood draws, diaper changes, wound care, ward rounds*” (physiotherapist 23). Both occupational therapists and physiotherapists noted that patients have the most energy in the morning, with lower energy and more pain in the afternoon after earlier exertion. As a result, most treatment is scheduled in the morning. Documentation is sometimes postponed until the afternoon, when monodisciplinary meetings also take place. Overall, there is significantly less physical activity in the afternoon. Physiotherapists also expressed the need to reserve time in the afternoon for acute or newly admitted patients. Nursing staff were not always aware of the physiotherapy offering later in the day and regretted that coverage from other professions was not the same in the afternoon. “*After all, patients are admitted around the clock*,” as Nurse 16 put it.

### Implications for cultural models of care

The absence of formalized interprofessional structures is not just an administrative gap; it reflects a model of care in which coordination is seen as optional rather than essential. The lack of clarity in interprofessional dialogue reflects competing cultural models embedded in organizational structures, which constrain opportunities for coordinated planning around mobilization. Similarly, pain management practices are sustained by a cultural expectation that patients must self-advocate, and that nurses, rather than the interprofessional team, hold primary responsibility for pain relief. This model overlooks the vulnerability of postoperative patients and the interdependency between professions. Finally, resource constraints are filtered through institutional logics that prioritize morning activity and minimize interprofessional presence in the afternoon, reinforcing a routine where mobilization becomes both time-bound and profession-bound.

### Mediating artefacts

From a cultural model perspective, the material environment and its artefacts are never neutral support for care. Following Hasse (44), artifacts are material or symbolic tool that are themselves shaped by cultural models, and that, in turn, reproduce them. They mediate human practices by making certain actions and understandings appear natural and legitimate, while rendering others unlikely or invisible. In this way, physical spaces and assistive devices are not just backdrops to mobilization but active mediators that embed professional routines, shape expectations of patient activity, and influence whether mobilization is seen as a therapeutic goal or a logistical task.

### Physical spaces as mediating artefacts

Distinct uniforms, keycard colors, and spatial arrangements, can be understood as cultural artefacts that embody and reproduce cultural models of professional identity and boundaries. The physiotherapists’ office, marked “Physio/OT” and often kept closed, not only defines a physical space but materializes a cultural model of ownership and autonomy over specialized knowledge. The expectation that the door should remain closed functions as a mediating artefact: it sustains a division of labor by protecting disciplinary space, while simultaneously constraining possibilities for cross-professional collaboration. In this sense, materiality is not neutral but actively mediates practice, subjectivity, and the reproduction of institutional boundaries.

Both wards have common rooms intended for social interaction, communal dining, or light activity. In practice, patients rarely use these spaces, and staff often repurpose them for storage or overflow. Design choices made without user input result in environments that staff perceive as uninviting or impractical.

Corridors, although equipped with handrails, are primarily occupied by trolleys, carts, and bed transports rather than patients. Even therapy rooms, spaces that could signal active rehabilitation, are shared with administrative work, sending mixed messages.: “*I think the signal it sends to patients is, ‘Am I going into an office to train?*’” (Physiotherapist 3).

These spatial arrangements exemplify how artefacts reinforce a cultural model that subordinates patient spaces to staff needs and positions the bed as the patients’ “default” location. Meals, entertainment, and personal items delivered bedside, reduce both opportunities and incentives for movement, embedding immobility into daily routines.

### Assistive devices as mediating artefacts

The Sara Stedy exemplifies how artefacts not only support but also redefine mobilization practice. Designed for patients unable to walk but able to stand, it is clinically valuable in cases such as epidural-induced numbness. Yet its widespread, habitual use extended far beyond these indications. Staff admitted it was often chosen for speed and safety during busy shifts, even when walking with assistance was possible:

> *"When I think back to the time before the Sara Stedy, for example, had even been invented, we just did it. We walked with them. And if they couldn’t, then they had to sit down again, and we had to figure out why they couldn’t. And if it was because of pain, then they had to be given some pain relief. But if they could walk, then they had to walk. Unfortunately, that practice has been significantly reduced*” (Physiotherapist 22).

As a mediating artefact, the Sara Stedy reflects and reinforces a shift in the cultural model of mobilization from an active, therapeutic process aimed at regaining walking ability, to a transfer task prioritized for efficiency and safety. In this sense, the artefact embodies and reproduces priorities of logistical flow, staff workload, and patient comfort, sometimes at the expense of rehabilitation goals.

### Implications for cultural models of care

Both physical spaces and assistive devices demonstrate how mediating artefacts shape, stabilize, and sometimes constrain cultural models. By embedding certain professional norms, such as efficiency over walking, these artefacts contribute to a cultural model in which mobilization is clinically valued yet operationally deprioritized. Recognizing artefacts as active mediators makes visible how cultural models are performed in practice and opens possibilities for reconfiguring material arrangement to better promote activity.

## DISCUSSION

The aim of this ethnographic study was to explore clinical practice related to mobilization, basic mobility, physical activity and exercise for patients during hospitalization following hip fracture surgery. Drawing on a critical-realist and hermeneutic framework, we sought to understand not only what professionals do, but how structural and cultural mechanisms shape their everyday practices.

We found that mobilization practices after hip fracture surgery is influenced by the interactions of divergent cultural models, organizational routines, and mediating artefacts. Difference in how professional groups define and prioritize mobilization, exercise, and physical activity, lead to unclear task boundaries and diffusion of responsibility, summarized by one physiotherapist as “*every man and no man’s responsibility*.” These mechanisms reinforced a predominantly mono-professional culture, in which physiotherapists are positioned as the primary drivers of mobilization, while interprofessional collaboration tends to unfold in parallel rather than integrated forms. In addition, pain management procedures, resource constraints and the physical environment further reflected cultural models that prioritized efficiency and logistical flow over sustained patient activity.

Using a critical-realist lens, we interpret these findings as evidence that the persistent knowing–doing gap in postoperative mobilization is not merely a matter of individual behavior or workload but emerges from underlying cultural and structural conditions that shape what forms of mobilization are perceived as feasible and legitimate.

### Interpretation in relation to existing literature

#### What are we talking about—and who is responsible for it?

While key terminology was defined at the outset, our empirical data reveal that mobilization remains the most dominant, and variably interpreted concept in clinical practice. Nurses tended to integrate mobilization within routine care activities (e.g., transferring patients to chairs or toilets), while physiotherapists reserved exercise for structured therapeutic interventions and used physical activity mainly in formal discussions of treatment goals. Similar, physiotherapy planning and nursing execution have been reported elsewhere (45), and a systematic review confirm that despite rhetorical support for mobility, in-hospital physical activity remains a low clinical priority (46). These differences reflect not just communication challenges, but deeper professional orientations formed through education and socialization. From the perspective of *relational agency* (47), nurses and physiotherapists operate with distinct *motive orientations*: nursing grounded in immediate care and task completion, physiotherapy focused on progressive functional recovery. Without a shared language or mutual understanding of goals, interprofessional work easily defaults to parallel practice—coexistence without integration. Such tendencies in interprofessional work reflect not only interpersonal barriers but also the organizational factors under which collaboration takes place.

#### Organizational factors

Interprofessional collaboration and coordinated care are widely recommended as standards for post-operative management of patients sustaining a hip fracture (48,49). Yet, our findings reveal an interprofessional collaboration pattern characterized by fragmented, task-oriented interactions, with few formalized structures and little continuity. This echoes Assafi et al. (50), who described how care in the same context was divided into specialized tasks that undermined collective responsibility and hindered coordination required for effective mobilization. They emphasized the need for physical presence and clearer work procedures to strengthen collaboration. Similarly, Stutzbach et al. (51), concluded that successful in-hospital mobility depends on effective interdisciplinary teamwork - when absent, mobilization becomes delayed, inconsistent, or deprioritized.

While existing literature often frames these barriers in logistical terms, our findings suggest that underlying cultural models play an equally important role. This becomes visible in pain management. Although adequate analgesia is a well-established prerequisite for mobilization (52), and requires coordination between nurses, physiotherapists, and pharmacy staff (53), our findings show that the procedure was shaped by unspoken role assumptions. Pain management was culturally defined as a nursing task, yet its initiation was frequently expected from the patients themselves. This mismatch reflects deeper procedural inefficiency, delaying mobilization because pain control was not collectively owned. Addressing this, requires explicit interprofessional dialogue to clarify how fundamental enablers of mobilization, such as analgesia, are defined, executed, and shared across roles.

A second organizational mechanism concerns resource allocation. Consistent with previous studies (46,54) staff reported insufficient personnel for two-person mobilizations. However, our findings add nuance by showing that constraints were largely temporal and structural. Therapists were primarily present during daytime hours, with most treatment clustered in the late morning, partly due to assumptions about patient fatigue in the afternoon. Documentation and new admissions dominated later in the day, leaving fewer opportunities for therapy. Yet, feasibility studies (10,55) and efficacy trials (11,56) have shown that afternoon and weekend rehabilitation are both achievable and effective when supported by organizational planning and workflow adjustments (56).

Thus, the issue is less about absolute staffing levels than about how time and professional resources are organized and sequenced within the daily rhythm of the ward. Resource allocation is not a neutral or purely logistical matter. It is shaped by professional boundaries, implicit hierarchies, and experiences about when mobilization is appropriate or realistic. These cultural models influence which tasks are prioritized, how time is valued, and which activities are perceived as feasible within a shift. As the number of older adults sustaining hip fractures continues to rise (57), optimizing mobility outcomes will require more than simply increasing staff numbers. It calls for a reconfiguration of workflows and a critical reflection on established routines, ensuring that time, space, and professional expertise are aligned around the shared goal of maintaining patient mobility.

### Mediating artefacts

The hospital bed has long been identified as a central artefact shaping both clinical practice and patient behavior (51). It symbolizes both safety and care, but simultaneously reinforces a cultural model in which passivity and immobility are normalized during hospitalization. In this way, the physical environment, including bed, layout, and surrounding routines, actively constrains opportunities for movement and may limit the fulfilment of professional goals aimed at supporting mobility.

A parallel shift in practice was evident in the growing reliance on assistive devices such as the Sara Stedy. Although staff universally acknowledged that walking independently is preferable to passive transfer, use of the device has increased over time, especially under conditions of high workload or limited staffing. What began as a pragmatic solution has gradually become institutionalized and socially accepted as "good enough" mobilization. This development can be understood through the lens of the Normalization Process Theory that explains how new practices become embedded and taken for granted in everyday clinical work (58). A routine stabilized not because it is clinically optimal, but because it aligns with prevailing organizational pressures and cultural logics.

Such normalization processes illustrate how artefacts, although originally designed to support care and enhance safety, can come to reflect and reinforce cultural models that unintentionally deprioritize patient activity. Importantly, this normalization may not only stem from logistical constraints but also from implicit risk assessments: concerns about patient safety and fear or falls further encourage the choice of safer, less active forms of mobility, even for patients capable of walking. These dynamics raises important questions for clinical practice: How do artefacts like the Sara Stedy come to define what counts as mobilization? And how might teams engage in reflective dialogue about when these artefacts enable, and when they constrain, patient autonomy and mobility*?*

### Strengths & limitations

A key strength of this study, supporting its credibility, is the alignment between study design, data collection, and research aim. The combination of participant observations and subsequent interviews allowed for analytical triangulation, where interview data reinforced and nuanced the preliminary interpretations from the field. Credibility was further enhanced through the use of direct quotations from transcribed material, providing transparency and illustrating how themes and subthemes were grounded in the empirical data (33).

Prior to the study start, healthcare professionals were informed about the study’s aim and that researchers would be present to observe clinical practice over the coming period. While such awareness may have influenced behavior, the study was conducted at one of Denmark’s largest university hospitals, where clinical research is a routine and accepted part of daily operations. Thus, any observer effect is likely to have been limited.

To ensure the trustworthiness of the findings, the qualitative content analysis was conducted systematically and with ongoing methodological reflection. The process was marked by iterative learning and close collaboration within the research team, where codes, patterns, and interpretations were continuously discussed, challenged, and refined. Final themes were developed through team-based consensus, ensuring that the analysis was both empirically grounded and critically robust. This collective approach strengthened the analytical process, from coding and de-/re-contextualization to abstraction and interpretative synthesis (42).

The present study focused on the perspectives of healthcare professionals and management, and did not include patients. To provide a more comprehensive understanding, interviews with patients and caregivers have also been conducted and will be published separately. Combined, the ethnographic fieldwork, interviews with staff, and forthcoming patient and caregiver perspectives will contribute a rich, in-depth account of the clinical practice and culture surrounding mobilization.

In line with the study’s interpretive approach, it is important to acknowledge the influence of researcher positionality. Most members of the research team are trained physiotherapists, and this professional background shaped how clinical practices were observed, how interview questions were framed, and how meaning was constructed during analysis. While disciplinary expertise enabled in-depth engagement with the topic, it may also have introduced specific lenses of interpretation. Ongoing reflexivity and interprofessional dialogue within the team were therefore essential to critically examine and balance these perspectives.

Finally, while the study emphasizes cultural models of mobilization, it is important to acknowledge that other conceptual frameworks may also illuminate the complexity of mobilization practices in acute hospital settings. For example, the socio-ecological model (59) highlights how behavior is shaped by dynamic interactions across individual, interpersonal, organizational, community, and policy levels. Future research may benefit from applying this framework to capture additional contextual layers influencing mobilization practices that were beyond the scope of this study.

## Conclusion

Our study contributes to an understanding of why mobilization remains inconsistently practiced despite guidelines, showing that these inconsistencies are deeply rooted in divergent cultural models of professional responsibility, patient agency, and organizational priorities. Moreover, our findings demonstrates that artefacts, institutional structures, and the material environment do not merely support care, but actively shape how mobilization and mobility are understood, performed and prioritized in everyday practice. This represent a shift in the perspective from asking “Why don’t health care professionals mobilize patients?” to “How is mobilization shaped by cultural models and the system they work within?”

### Implications for clinical practice

To support clinical practice in meeting recommended levels of mobilization, physical activity, and exercise during acute hospitalization following hip fracture surgery, the findings of this study point to several key areas for improvement.

1. *Interprofessional collaboration as a key determinant* Establish a shared understanding across professional groups—not only of each other’s roles and priorities, but also of the joint responsibility for mobilization. Strengthen relational agency by aligning terminology and clarifying responsibilities to enable coordinated, interprofessional action.
2. *Adapt mobilization guidelines to fit your interprofessional practice* International guidelines clearly emphasize early weight-bearing and mobilization postoperatively, but offer limited guidance on the type, intensity, frequency, and duration of subsequent activity. Local adaptation is essential—interpret and discuss guidelines within the interprofessional team to ensure they align with patient needs and contextual realities.
3. *Resource availability and allocation* Address not just how many resources are available, but how they are distributed throughout the day and week. Reflect on current priorities: Can workflows be optimized within existing capacity, or are additional resources necessary to meet guideline recommendations?
4. *Physical environment as a facilitator* Involve both staff and patients in shaping physical spaces that promote movement. The material environment does more than motivate—it can act as a mediating artefact that reinforces a cultural model supporting active recovery.
5. *Maintain curiosity about your own practice* Mobilization in an orthopedic ward may appear straightforward but is shaped by complex interprofessional dynamics, cultural norms, and systemic constraints. Continuous reflection can uncover opportunities for improvement and innovation.

## Data Availability

Data are not available.

## Declarations of interest

None

## Funding source

This work was supported by grants from the Association of Danish Physiotherapists and the Research Fund of Copenhagen University Hospital, Amager and Hvidovre, Hvidovre, Denmark. The funding sources had no role in this work.

## CRediT authorship statement

**Maria Swennergren Hansen:** Conceptualization, Formal analysis, Funding acquisition, Investigation, Methodology, Project administration, Supervision, Validation, Writing - original draft and review & editing. **Meta Ellen Staal Høite:** Conceptualization, Formal analysis, Investigation, Validation, Writing - review & editing. **Thomas Bandholm:** Conceptualization, Formal analysis, Funding acquisition, Validation, Writing - review & editing. **Sofie Tscherning Lindholm:** Formal analysis, Funding acquisition, Methodology, Validation, Writing - review & editing. **Kira Marie Skibdal:** Conceptualization, Formal analysis, Investigation, Methodology, Validation, Writing - review & editing. **Mette Merete Pedersen:** Conceptualization, Formal analysis, Funding acquisition, Investigation, Methodology, Supervision, Validation, Writing - review & editing. **Jeanette Wassar Kirk:** Conceptualization, Formal analysis, Funding acquisition, Investigation, Methodology, Supervision, Validation, Writing - original draft and review & editing.

## Acknowledgements

We are grateful for the support and help from staff, and management in the participating wards.

## Declaration of generative AI and AI-assisted technologies in the manuscript preparation process

During the preparation of this work the authors used ChatGPT in order to improve the readability of the text. After using this tool, the authors reviewed and edited the content as needed and take full responsibility for the content of the published article.

## Notes

### Competing Interest Statement

The authors have declared no competing interest.

### Author Declarations

The study was approved by The Research Ethics Committee of the Capital Region of Denmark (journal number: F-23067628, 8/11/2023)

## REFERENCES

1. Kristensen PK, Röck ND, Christensen HC, Pedersen AB. The danish multidisciplinary hip fracture registry 13-year results from a population-based cohort of hip fracture patients. Clin Epidemiol. 2020;12:9–21.

2. Yan B, Sun W, Wang W, Wu J, Wang G, Dou Q. Prognostic significance of frailty in older patients with hip fracture: a systematic review and meta-analysis. Int Orthop. 2022 Dec 1;46(12):2939–52.

3. Zidén L, Wenestam CG, Hansson-Scherman M. A life-breaking event: Early experiences of the consequences of a hip fracture for elderly people. Clin Rehabil. 2008;22(9):801–11.

4. Abrahamsen C, Viberg B, Nørgaard B. Patients’ perspectives on everyday life after hip fracture: A longitudinal interview study. Int J Orthop Trauma Nurs. 2022 Feb 1;44.

5. Morri M, Ambrosi E, Chiari P, Orlandi Magli A, Gazineo D, D’ Alessandro F, et al. One-year mortality after hip fracture surgery and prognostic factors: a prospective cohort study. Sci Rep. 2019 Dec 1;9(1).

6. Teasell R, Dittmer DK. Complications of Immobilization and Bed Rest Part 2: Other complications. Vol. 39, Canadian Family Physician. 1993.

7. Tantigate D, Jansatjawan N, Adulkasem N, Ramart P, Riansuwan K. Risk factors for postoperative urinary retention in fragility hip fracture patients: a prospective study. BMC Geriatr. 2024 Dec 1;24(1).

8. Tarazona-Santabalbina FJ, Belenguer-Varea Á, Rovira E, Cuesta-Peredó D. Orthogeriatric care: Improving patient outcomes. Clin Interv Aging. 2016 Jun 24;11:843–56.

9. Agarwal N, Feng T, Maclullich A, Duckworth A, Clement N. Early mobilisation after hip fracture surgery is associated with improved patient outcomes: A systematic review and meta-analysis. Musculoskeletal Care. 2024 Mar 1;22(1).

10. Zilmer CK, Kristensen MT, Magnusson SP, Bährentz IB, Jensen TG, Zoffmann SØ, et al. Intensified acute in-hospital physiotherapy for patients after hip fracture surgery: a pragmatic, randomized, controlled feasibility trial. Disabil Rehabil. 2023 Dec;1–10.

11. Kimmel LA, Liew SM, Sayer JM, Holland AE. HIP4Hips (High intensity physiotherapy for hip fractures in the acute hospital setting): A randomised controlled trial. Medical Journal of Australia. 2016 Jul 18;205(2):73–8.

12. March MK, Dennis SM, Caruana S, Mahony C, Elliott JM, Polley S, et al. Boosting inpatient exercise after hip fracture using an alternative workforce: a mixed methods implementation evaluation. BMC Geriatr. 2024 Dec 1;24(1).

13. Kristensen MT, Öztürk B, Röck NDi, Ingeman A, Palm H, Pedersen AB. Regaining pre-fracture basic mobility status after hip fracture and association with post-discharge mortality and readmission - A nationwide register study in Denmark. Age Ageing. 2019 Mar 1;48(2):278–84.

14. Zusman EZ, Dawes MG, Edwards N, Ashe MC. A systematic review of evidence for older adults’ sedentary behavior and physical activity after hip fracture. Clin Rehabil. 2018 May 1;32(5):679–91.

15. Hansen MS, Kristensen MT, Zilmer CK, Berger AL, Kirk JW, Marie Skibdal K, et al. Very low levels of physical activity among patients hospitalized following hip fracture surgery: a prospective cohort study. Disabil Rehabil [Internet]. 2025 Jan 21;1–10.

16. Kronborg L, Bandholm T, Palm H, Kehlet H, Kristensen MT. Physical activity in the acute ward following hip fracture surgery is associated with less fear of falling. J Aging Phys Act. 2016 Oct 1;24(4):525–32.

17. Kirk JW, Bodilsen AC, Sivertsen DM, Husted RS, Nilsen P, Tjørnhøj-Thomsen T. Disentangling the complexity of mobility of older medical patients in routine practice: An ethnographic study in Denmark. PLoS One. 2019 Apr 1;14(4).

18. Westerlund A, Sundberg L, Nilsen P. Implementation of Implementation Science Knowledge: The Research-Practice Gap Paradox. Vol. 16, Worldviews on Evidence-Based Nursing. Blackwell Publishing Ltd; 2019. p. 332–4.

19. Kristensen N, Nymann C, Konradsen H. Implementing research results in clinical practice- the experiences of healthcare professionals. BMC Health Serv Res. 2016 Feb 10;16(1).

20. Johansen LK, Larsen TS, Kirk JW, Pedersen BS, Nielsen BR, Kallemose T, et al. Exploring in-hospital mobility practices for geriatric patients: Insights from a mixed-method study. BMC Geriatr. 2025 May 13;25(1):330.

21. Paes VM, Ting A, Masters J, Paes MVI, Graham SM, Costa ML. A systematic review of evidence regarding the association between time to mobilization following hip fracture surgery and patient outcomes. Bone Jt Open. 2025 Jun;(7):741–7.

22. Archer M. Critical Realism: Essential Readings. London: Routledge; 1998.

23. Buch-Hansen H, Nielsen P. Kritisk realisme . Frederiksberg: Roskilde Universitetsforlag; 2005.

24. Geanellos R. Exploring Ricoeur’s hermeneutic theory of interpretation as a method of analysing research texts. Nurs Inq. 2000;7(2):112–9.

25. Gillo MD. Fundamentals of Hermeneutics as A Qualitative Research Theoretical Framework. European Journal of Education and Pedagogy. 2021 Jun 20;2(3):42–5.

26. Hoyer EH, Brotman DJ, Chan KS, Needham DM. Barriers to early mobility of hospitalized general medicine patients: Survey development and results. Am J Phys Med Rehabil. 2015 Apr 20;94(4):304–12.

27. World Health Organization. Physical activity [Internet]. [cited 2025 Dec 15]. Available from: https://www.who.int/news-room/fact-sheets/detail/physical-activity

28. Exercise Therapy - MeSH - NCBI [Internet]. [cited 2025 Dec 15]. Available from: https://www.ncbi.nlm.nih.gov/mesh/68005081

29. Kristensen MT, Andersen L, Bech-Jensen R, Moos M, Hovmand B, Ekdahl C, et al. High intertester reliability of the Cumulated Ambulation Score for the evaluation of basic mobility in patients with hip fracture. Clin Rehabil. 2009 Dec;23(12):1116–23.

30. Kristensen MT, Turabi R, Sheehan KJ. The relationship between extent of mobilisation within the first postoperative day and 30-day mortality after hip fracture surgery. Clin Rehabil. 2024 Jul;38(7):990–997.

31. Spradley JP. Participant observation. New York: Holt, Rinehart and Winston; 1980.

32. Hammersley M, Atkinson P. Ethnography: Principles in practice. Third edition. London: Routledge; 2007.

33. O’Brien BC, Harris IB, Beckman TJ, Reed DA, Cook DA. Standards for reporting qualitative research: A synthesis of recommendations. Academic Medicine. 2014;89(9):1245–51.

34. Turnock C, Gibson V. Validity in action research: A discussion on theoretical and practice issues encountered whilst using observation to collect data. J Adv Nurs. 2001 Nov;36(3):471–7.

35. Tracy SJ. Qualitative quality: Eight a"big-tent" criteria for excellent qualitative research. Qualitative Inquiry. 2010 Dec;16(10):837–51.

36. Etikan I. Comparison of Convenience Sampling and Purposive Sampling. American Journal of Theoretical and Applied Statistics. 2016;5(1):1.

37. Association WM. World Medical Association Declaration of Helsinki: Ethical Principles for Medical Research Involving Human Subjects. JAMA. 2013 Nov 27;310(20):2191–4.

38. Tjørnhøj-Thomsen T. Samværet. Tilblivelse i tid og rum. In: Hastrup K, editor. Ind i verden En grundbog i antropologisk metode. København: Hans Reitzels Forlag; 2010. p. 93–117.

39. Bandholm T, Thorborg K, Ardern CL, Christensen R, Henriksen M. Writing up your clinical trial report for a scientific journal: the REPORT trial guide for effective and transparent research reporting without spin. Br J Sports Med. 2022;

40. Kvale S, Brinkmann S. Interview - Det kvalitative forskningsinterview som håndværk. 3rd ed. Copenhagen: Hans Reizels Forlag; 2015.

41. Graneheim UH, Lundman B. Qualitative content analysis in nursing research: Concepts, procedures and measures to achieve trustworthiness. Nurse Educ Today. 2004;24(2):105–12.

42. Lindgren BM, Lundman B, Graneheim UH. Abstraction and interpretation during the qualitative content analysis process. Int J Nurs Stud. 2020 Aug 1;108.

43. Hasse C. Kulturanalyse i organisationer. Begreber, metoder og forbløffende læreprocesser. 2nd ed. Vol. 1. Frederiksberg: Samfundslitteratur; 2011. 96–99 p.

44. Hasse C. An Anthropology of Learning: On Nested Frictions in Cultural Ecologies. New York : Springer Berlin Heidelberg; 2014.

45. Haslam-Larmer L, Donnelly C, Auais M, Woo K, DePaul V. Early mobility after fragility hip fracture: a mixed methods embedded case study. BMC Geriatr. 2021 Dec 1;21(1).

46. Alsop T, Woodforde J, Rosbergen I, Mahendran N, Brauer S, Gomersall S. Perspectives of health professionals on physical activity and sedentary behaviour in hospitalised adults: A systematic review and thematic synthesis. Clin Rehabil. 2023 Oct 1;37(10):1386–405.

47. Edwards A. The 43rd Vernon-Wall Lecture: What working relationally brings to problem-solving. British Journal of Educational Psychology. 2025;

48. Drew S, Fox F, Gregson CL, Gooberman-Hill R. Model of multidisciplinary teamwork in hip fracture care: A qualitative interview study. BMJ Open. 2024 Feb 27;14(2).

49. Riemen AHK, Hutchison JD. The multidisciplinary management of hip fractures in older patients. Orthop Trauma. 2016 Apr 1;30(2):117–22.

50. Assafi L, Evaristi D, Trevino CS, Larsen TS. It’s all about presence: Health professionals’ experience of interprofessional collaboration when mobilizing patients with hip fractures. J Interprof Care. 2021;

51. Stutzbach J, Jones J, Taber A, Recicar J, Burke RE, Stevens-Lapsley J. Systems Approach Is Needed for In-Hospital Mobility: A Qualitative Metasynthesis of Patient and Clinician Perspectives. Arch Phys Med Rehabil. 2021 May 1;102(5):984–98.

52. Münter KH, Clemmesen CG, Foss NB, Palm H, Kristensen MT. Fatigue and pain limit independent mobility and physiotherapy after hip fracture surgery. Disabil Rehabil. 2018 Jul 17;40(15):1808–16.

53. McDonough CM, Harris-Hayes M, Kristensen MT, Overgaard JA, Herring T, Kenny AM, et al. Physical therapy management of older adults with hip fracture. Journal of Orthopaedic and Sports Physical Therapy. 2021 Feb 1;51(2):CPG1–81.

54. Brown CJ, Williams BR, Woodby LL, Davis LL, Allman RM. Barriers to mobility during hospitalization from the perspectives of older patients and their nurses and physicians. J Hosp Med. 2007 Sep;2(5):305–13.

55. Lau B, March MK, Harmer AR, Caruana S, Mahony C, Dennis S. Experiences of Boosting Inpatient Exercise After HipFracture Surgery Using An Alternative Workforce - A Qualitative Study. BMC Geriatr. 2024 Dec 1;24(1).

56. Ogawa T, Onuma R, Sagae H, Schermann H, Kristensen MT, Fushimi K, et al. Association between additional weekend rehabilitation and functional outcomes in patients with hip fractures: Does age affect the effectiveness of weekend rehabilitation? Eur Geriatr Med. 2024 Aug;15(4):1091–1100.

57. Sing C. Global epidemiology of hip fractures: secular trends in incidence rate, post-fracture treatment, and all-cause mortality. J Bone Miner Res. 2023 Aug;38:1064–75.

58. May CR, Mair F, Finch T, MacFarlane A, Dowrick C, Treweek S, et al. Development of a theory of implementation and integration: Normalization Process Theory. Implementation Science. 2009;4(1).

59. Mcleroy KR, Bibeau D, Steckler A, Glanz K. An Ecological Perspective on Health Promotion Programs. 1988;15(4):351–77. Available from: https://www.jstor.org/stable/45049276

